# Association of Oral Cancer site with addiction and Sociodemographic characteristics: A Cross sectional study Conducted at a Tertiary Health Centre

**DOI:** 10.1101/2024.02.20.24303029

**Authors:** Praful Maroti Hulke, Jalindhar Pandurang Baravakar, Vinod Genu Bagade, Alka Modi Asati

## Abstract

**Introduction:** In contemporary epidemic scenarios, oral cancer ranks within the top 3 cancer types afflicting the Indian population. The primary risk factors include alcohol consumption, tobacco usage in various forms such as cigarettes and smokeless tobacco, betelnut chewing, and infection with the human papilloma virus.

**Materials and Methods:** This study, conducted at a tertiary healthcare centee, adopted a hospital-based cross-sectional approach involving 233 oral cancer patients who sought medical care from January 1st to December 31st, 2017.

**Results:** The findings indicate that males above the age of 60 (40.11%) and females aged between 41 and 50 years (45.45%) were the most affected groups, a statistically significant association (p<0.05) was found. The primary sites for oral cancer were identified as the cheek (40.36%) and tongue (31.78%). Remarkably, Majority of male patients exhibited a combination of addictions, including tobacco chewing, smoking, and alcohol use, whereas most female patients were exclusively involved in tobacco chewing.

**Conclusion:** In order to curb the escalating prevalence of oral cancer in India, it is imperative to implement comprehensive public education initiatives and health promotion strategies aimed at reducing both smoking and alcohol consumption.

## Introduction

In today’s world, non-communicable diseases, often called as modern epidemics, are on the rise. Cancer, the second leading cause of death in developed nations among the top 10 mortality causes, plays a significant role in this trend (1). Oral cancer, an ancient ailment documented in texts like the Sushruta Samhita, is a malignant neoplasm affecting various oral structures, including the lip, mouth floor, cheek lining, gingiva, palate, and tongue. Its local aggressiveness leads to disfigurement, functional impairment, and physical and psychosocial distress, ultimately affecting an individual’s quality of life (3). Globally, oral cancer ranks as the 6th most prevalent cancer, with notable geographical variations (4). In India, it constitutes nearly 40% of all cancer cases and remains among the top 3 cancer types (5). Alarmingly, over five individuals succumb to oral cancer every hour in India (7). Common risk factors for oral cancer include tobacco product usage, such as cigarettes, smokeless tobacco, betel nut chewing, excessive alcohol consumption, and human papillomavirus (HPV) infection. Low socio-economic status, often linked to factors like nutrition and personal habits, is also associated with an elevated risk (3).

India, a developing nation, faces a unique challenge 22% of its population living below the poverty line. A cancer diagnosis often exacerbates the economic struggles of affected families (8)(9). To monitor cancer incidence, survival, and mortality, the Indian Council of Medical Research initiated the National Cancer Registry Programme (NCRP) in 1982. This program includes population-based cancer registration (PBCR), with “Barshi” as the primary registry for rural areas in western Maharashtra and the Marathwada region. The incidence of cancer varies significantly across Indian cities, with rates ranging from 37.3 per 100,000 in Barshi to 86.7 per 100,000 in Chennai among males and 44.1 per 100,000 in Barshi to 101.2 per 100,000 in Chennai among females, according to Population-Based Cancer Registries (PBCRs). The estimated overall cancer incidence in India ranges from 70 to 90 cases per 100,000 population, resulting in approximately 7 to 9 lakh new cancer cases annually. In the year 2000, cancer was responsible for 5.5 lakh deaths in India, with Maharashtra accounting for 9% of these cases (10). It is crucial to dispel two misconceptions about cancer: first, that it is inevitable, and second, that it only affects industrialized nations. In reality, about 80% of cancer cases are preventable and linked to environmental factors (2). Early detection of oral cancer and mitigating risk factors offer the best chance of long-term survival, improved treatment outcomes, and more cost-effective healthcare.

With this perspective in mind, we conducted a study on oral cancer patients at a tertiary healthcare center to investigate the relationship between risk factors and oral cancer patients seeking care.

## Materials and Methods

### Study design

This hospital-based descriptive study, employing a cross-sectional design, was conducted on oral cancer patients.

### Place and duration of study

Patients were attending a tertiary healthcare centre over a one-year period from January 1, 2017, to December 31, 2017.

### Ethical approval and patient consent

It is an observational study; no intervention was made. Before commencement of this study – Institutional Ethical Committee “P.G. Coordination committee, Dr V. M. Govt. Medical College, Solapur, number 535, dated 06/09/2016” had approved this protocol.

### Inclusion and exclusion criteria

All diagnosed cases of oral cancer attending the Tertiary Health Care Centre during the study period who consented to participate were included. Out of 241 diagnosed oral cancer cases, 233 patients met the inclusion criteria.

### Sampling size and sampling

Patients were attending a tertiary healthcare center over a one-year period and who met the inclusion criteria were included in the study. So tenure sampling method was used.

### Data Collection

Permission was obtained from the surgery and ENT departments, and individuals were informed about the study. Informed written consent in the local language was obtained before participation. A separate proforma was completed for each patient and their relatives, ensuring patient identity remained confidential. A pre-designed, pre-tested proforma was used to gather information on socio-demographic characteristics, personal habits such as tobacco consumption (including chewing, betel quid, smoking, gutkha, khaini, or other forms), alcohol consumption (including duration, frequency, and amount), and the impact of these habits on living standards.

### Data Analysis

Statistical software was employed to analyze the data, which was presented in tables, figures, and graphs as needed. Descriptive and inferential analyses were performed using suitable hypothesis testing methods. Data analysis was carried out using SPSS software version 16.

## Results

This cross-sectional study, conducted over a one year from January 1, 2017, to December 31, 2017, included 233 diagnosed oral cancer patients attending a tertiary care center. The key findings are summarized below.

Table No. 1 displays oral cancer patients’ age and site-wise distribution by gender. The age range of affected individuals was 26 to 81 years, with the highest number of oral cancer patients in the above 60 age group, accounting for 32.61%, followed by 32.19% in the 51-60 age group. The least affected age group was below 30 years, at 1.29%. Gender-wise distribution revealed that 71.67% of patients were males, while 28.32% were females, resulting in a male-to-female ratio of 2.5:1. The most affected age group among males was above 60 years (40.11%), while among females, it was 41-50 years (45.45%). The difference in age and gender-wise distribution was statistically significant (p<0.05).

**Table No. 1:**
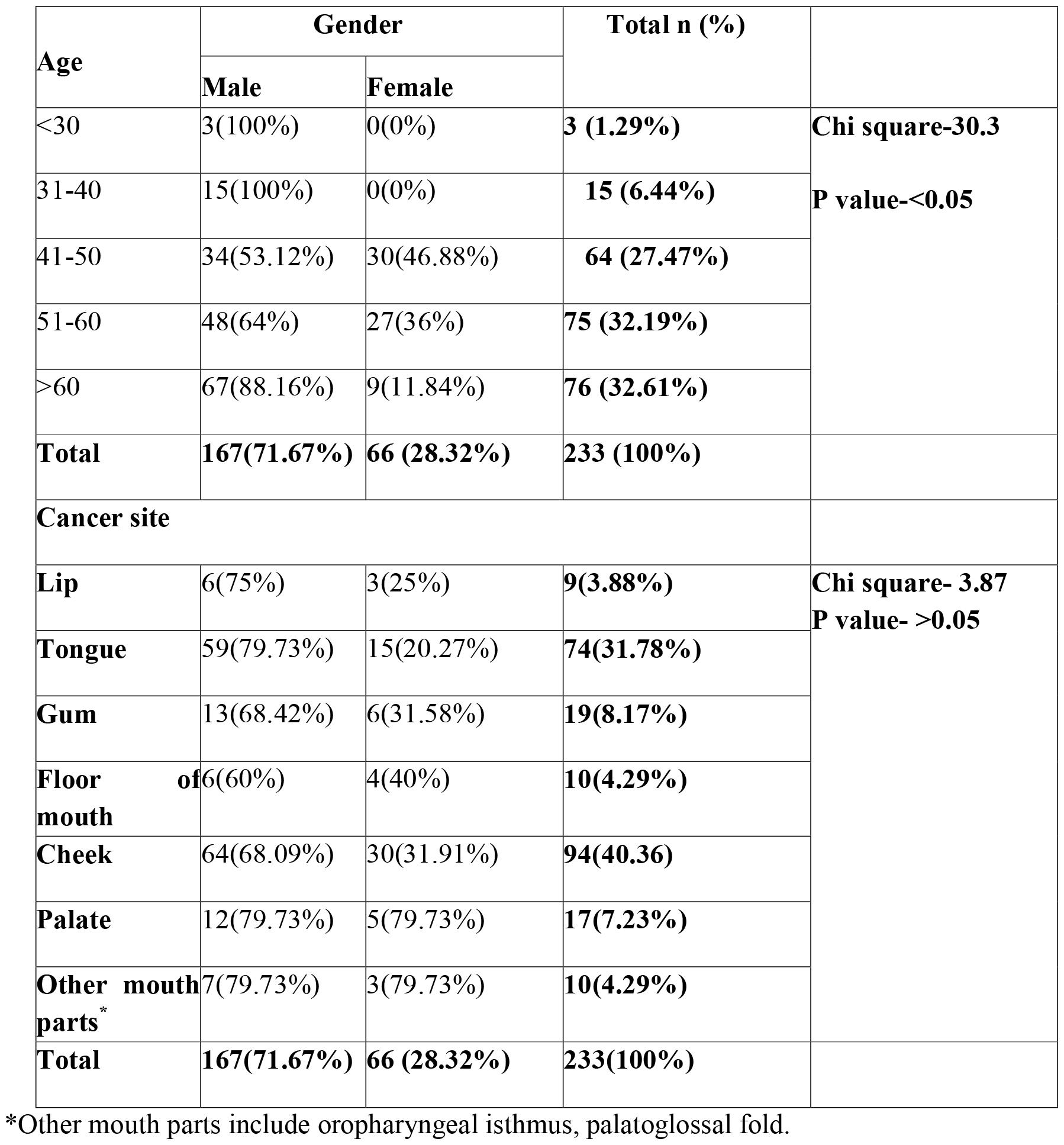
Distribution of patients according to age, site of cancer with gender(N=233):

| Age | Gender |  | Total n (%) |  |
| --- | --- | --- | --- | --- |
|  | Male | Female |  |  |
| <30 | 3(100%) | 0(0%) | 3 (1.29%) | Chi square-30.3<br>P value-<0.05 |
| 31-40 | 15(100%) | 0(0%) | 15 (6.44%) |  |
| 41-50 | 34(53.12%) | 30(46.88%) | 64 (27.47%) |  |
| 51-60 | 48(64%) | 27(36%) | 75 (32.19%) |  |
| >60 | 67(88.16%) | 9(11.84%) | 76 (32.61%) |  |
| **Total** | **167(71.67%)** | **66 (28.32%)** | **233 (100%)** |  |
| **Cancer site** |  |  |  |  |
| **Lip** | 6(75%) | 3(25%) | 9(3.88%) | Chi square- 3.87<br>P value- >0.05 |
| **Tongue** | 59(79.73%) | 15(20.27%) | 74(31.78%) |  |
| **Gum** | 13(68.42%) | 6(31.58%) | 19(8.17%) |  |
| **Floor of mouth** | 6(60%) | 4(40%) | 10(4.29%) |  |
| **Cheek** | 64(68.09%) | 30(31.91%) | 94(40.36) |  |
| **Palate** | 12(79.73%) | 5(79.73%) | 17(7.23%) |  |
| **Other mouth parts\*** | 7(79.73%) | 3(79.73%) | 10(4.29%) |  |
| **Total** | **167(71.67%)** | **66 (28.32%)** | **233(100%)** |  |

\*Other mouth parts include oropharyngeal isthmus, palatoglossal fold.

**Figure 1:**
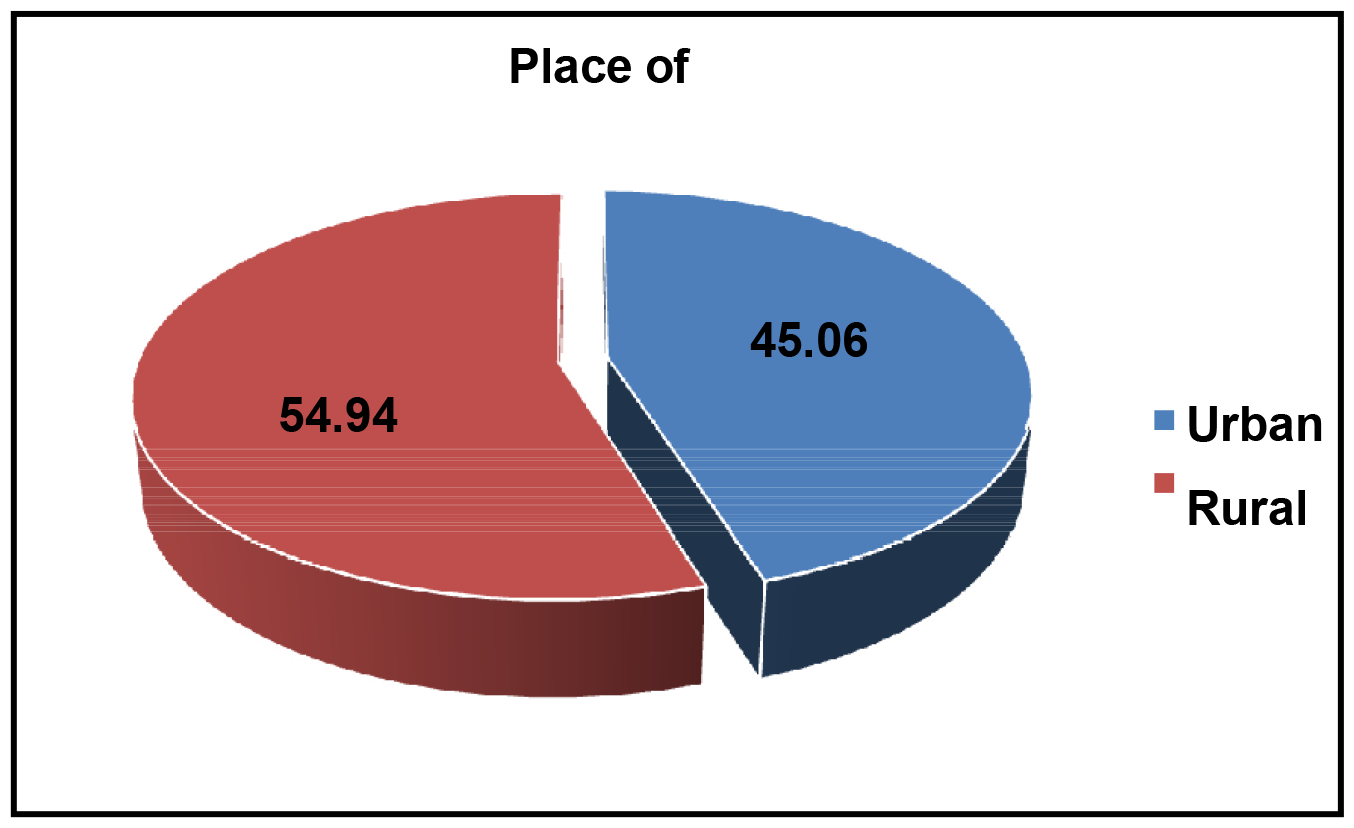
Distribution of patients according to Residence: [**Chi square value for goodness of fit is 2.27, p>0.05, not significant]** Figure No. 1 shows the distribution of patient according to their place of residence. Out of 233 oral cancer patients, 54.94% live in urban areas, and 45.06 % live in rural areas. This difference found was not statistically significant (p>0.05).

**Chart 1:**
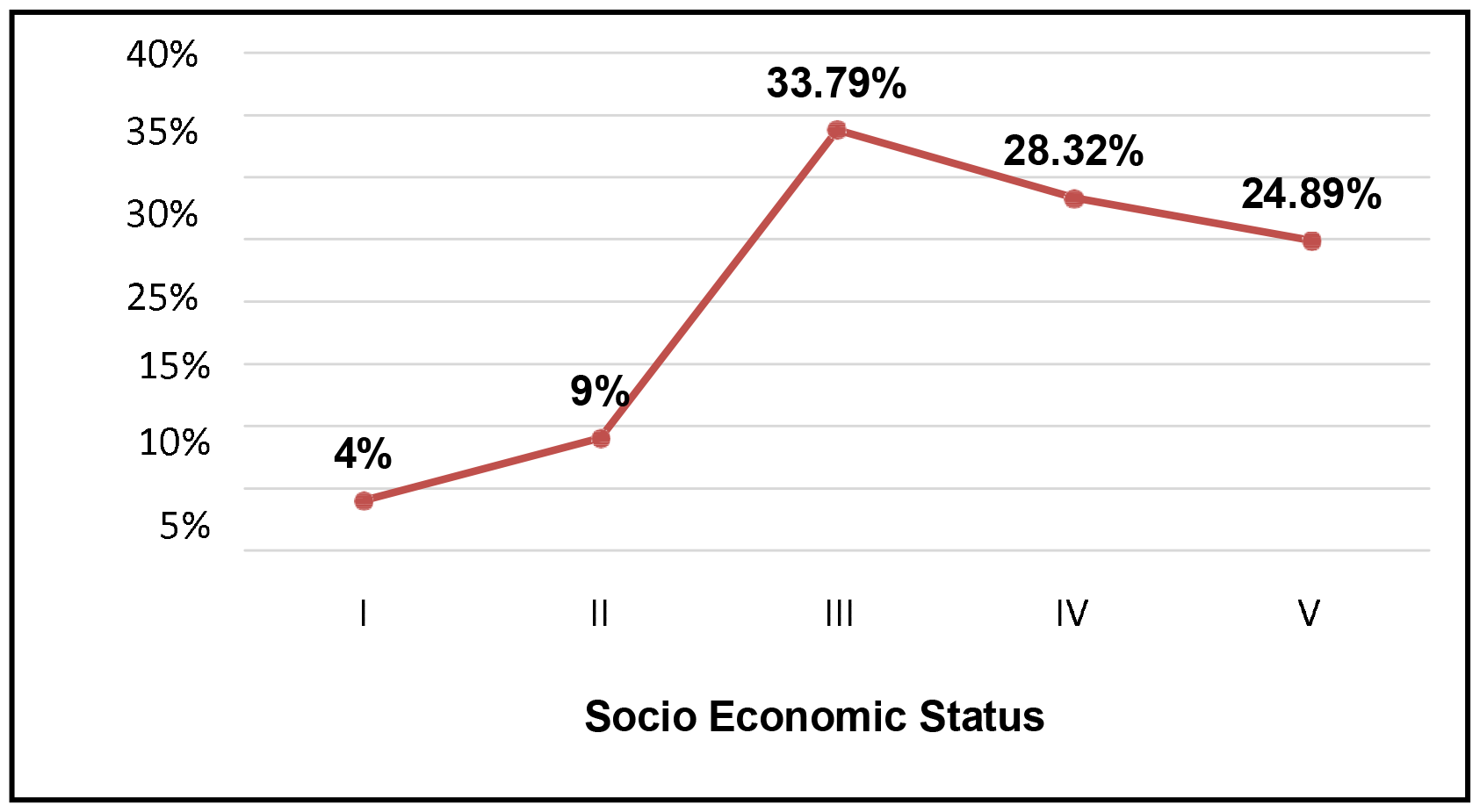
Distribution of patients according to Socio Economic Status: Chi-square=77.794. p <0.01

Figure No. 2 shows Regarding the distribution of patients by place of residence, out of 233 oral cancer patients, 54.94% lived in urban areas, and 45.06% lived in rural areas. However, this difference was not statistically significant (p>0.05). The majority of patients, 33.79%, belonged to class III socio-economic status (according to Modified B.G. Prasad’s classification), followed by 28.32% in class IV socio-economic status. The least affected patients belonged to class I socio-economic status. This difference was highly significant (p<0.01).

Table No. 2 illustrates the age and site-wise distribution of oral cancer patients. Cheek (40.36%) was the most common site of oral cancer, followed by the tongue (31.78%) and gum (8.17%), while the lip (3.88%) was the least affected site. The floor of the mouth and other mouth parts (oropharyngeal isthmus and palatoglossal fold) were equally affected (4.29%). No statistically significant association was found between cancer site and age at diagnosis (p>0.05).

**Table No. 2:** Age and cancer site-wise distribution of patients(N=233):

| Cancer site | Age at diagnosis (years) |  |  | Total No (%) |  |
| --- | --- | --- | --- | --- | --- |
|  | < 40 | 40 – 60 | >60 |  |  |
| **Lip** | 00 | 07(77.78%) | 02(22.22%) | **9(3.88%)** | **Chi-square-6.54**<br>**P value->0.05** |
| **Tongue** | 5(6.76%) | 43(58.11%) | 26(35.13%) | **74(31.78%)** |  |
| **Gum** | 02(10.53%) | 13(68.42%) | 04(21.05%) | **19(8.17%)** |  |
| **Floor of mouth** | 00 | 06(60%) | 04(40%) | **10(4.29%)** |  |
| **Cheek** | 7(11.45%) | 55(58.51%) | 32(30.04%) | **94(40.36%)** |  |
| **Palate** | 02(11.76%) | 10(58.82%) | 05(29.41%) | **17(7.23%)** |  |
| **Other mouth parts<sup>#</sup>** | 2(20%) | 5(50%) | 3(30%) | **10(4.29%)** |  |
| **Total** | **18(7.73%)** | **139(59.75%)** | **76(32.61%)** | **233(100%)** |  |

<sup>#</sup>Other mouth parts include oropharyngeal isthmus and palatoglossal fold.

Table No. 3 presents the distribution of patients by their habit patterns in about gender and cancer site. The majority of oral cancer patients had the habit of tobacco chewing alone (32.18%), followed by tobacco chewing combined with smoking (18.88%). Among males, the most common habit was tobacco chewing combined with alcohol and smoking (22.16%), followed by tobacco chewing combined with smoking (21.56%). Among females, tobacco chewing alone was more common (63.64%) than males (19.76%). This association between gender and habit patterns was highly significant (p<0.01). In terms of the distribution of patients according to their habit patterns and cancer site, the tongue was the common site of cancer when a single habit, be it chewing, smoking, or alcohol consumption, was present (56%). The cheek was the common site In cases with a combination of more than one habit. Patients with no habits were more likely to develop cancer in other mouth parts like the oropharyngeal isthmus and palatoglossal fold (40%). This association between cancer site and habit pattern was highly significant (p=0.00). Moreover, 54.94% of oral cancer patients had the habit for more than 10 years, while 36.05% had the habit for 5-10 years, and this difference was highly significant (p<0.01).

**Table No. 3:** Distribution of patients according to addiction with gender and cancer site (N=233):

| Addiction | Tobacco chewing alone | Smoking alone | Alcohol alone | Tobacco chewing + Smoking | Tobacco chewing + Alcohol | Smoking + Alcohol | Tobacco chewing + Alcohol + smoking | No Habits | Total |  |
| --- | --- | --- | --- | --- | --- | --- | --- | --- | --- | --- |
| **Male** | 33(19.76%) | 16(9.58%) | 11(6.59%) | 36(21.56%) | 10(5.99%) | 21(12.57%) | 37(22.16%) | 3(1.8%) | **167(71.67%)** | **p<0.01** |
| **Female** | 42(63.64%) | 3(4.55%) | 1(1.52%) | 8(12.12%) | 4(6.06%) | 2(3.03%) | 0 | 6(9.09%) | **66(28.32%)** |  |
| **Cancer site** |  |  |  |  |  |  |  |  |  |  |
| **Lip** | 9(100%) | 0 | 0 | 0 | 0 | 0 | 0 | 0 | **9(3.88%)** | **p=0.00** |
| **Tongue** | 42(56%) | 16(21.6%) | 10(13.51%) | 5(6.75%) | 0 | 0 | 0 | 1(1.35%) | **74(31.78%)** |  |
| **Gum** | 6(31.57%) | 0 | 0 | 12(63.15%) | 0 | 0 | 0 | 1(5.26%) | **19(8.17%)** |  |
| **Mouth** | 4(40%) | 0 | 0 | 6(60%) | 0 | 0 | 0 | 0 | **10(4.29%)** |  |
| **Cheek** | 14(14.89%) | 3(3.19%) | 2(2.12%) | 21(22.34%) | 14(14.89%) | 21(22.34%) | 19(20.21%) | 0 | **94(40.36%)** |  |
| **Palate** | 0 | 0 | 0 | 0 | 0 | 2(11.76%) | 12(70.58%) | 3(17.64%) | **17(7.23%)** |  |
| **Other\*** | 0 | 0 | 0 | 0 | 0 | 0 | 6(60%) | 4(40%) | **10(4.29%)** |  |
| **Total** | **75(32.18%)** | **19(8.15%)** | **12(5.15%)** | **44(18.88%)** | **14(6.04%)** | **23(9.87%)** | **37(15.87%)** | **9(3.86%)** | **233(100%)** |  |

\*Other mouth parts include oropharyngeal isthmus and palatoglossal fold.

**Figure No. 3:**
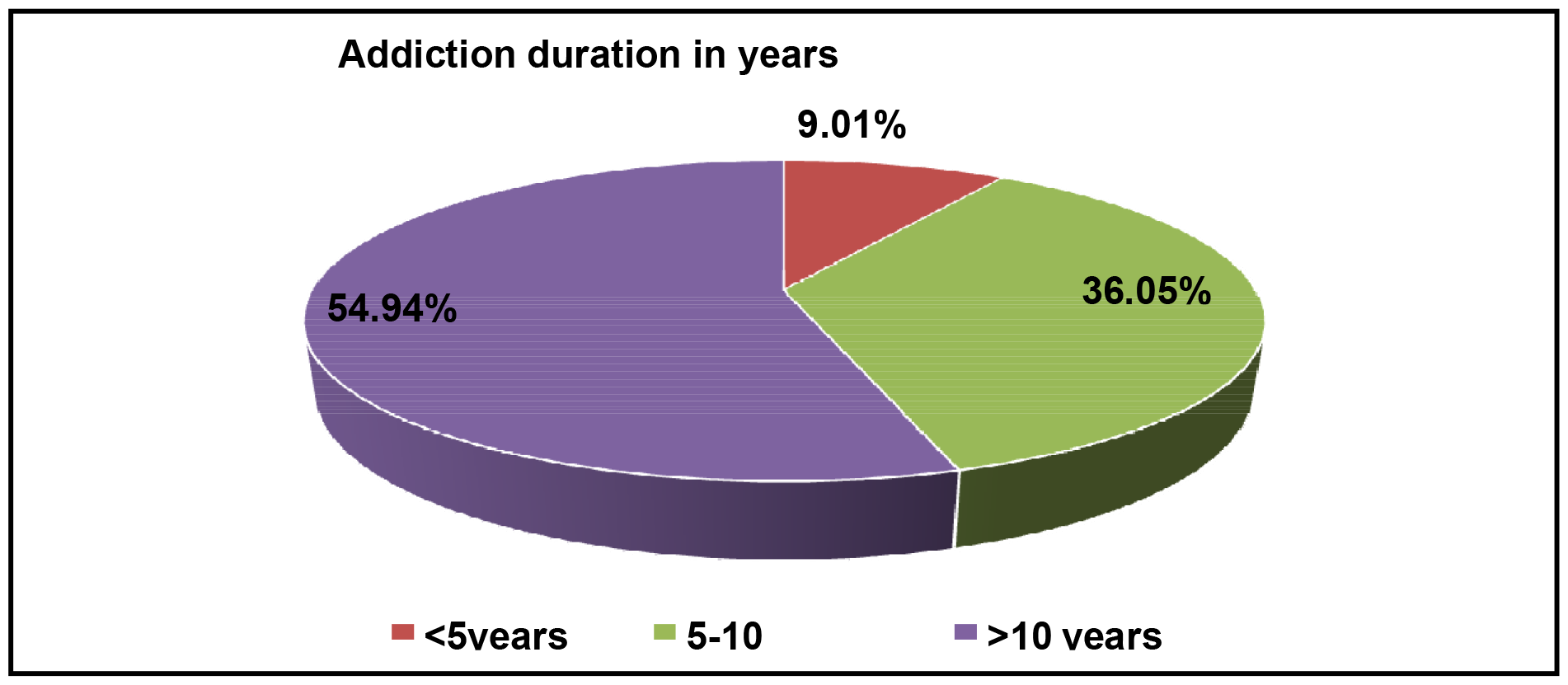
Distribution of patients according to duration of habit. [Chi square value for goodness of fit is 74.495, df=2,p<0.01, highly significant]

Figure No. 3 shows the distribution of patients according to their addiction duration. In the present study, it was seen that 54.94% of oral cancer patients had the habit for more than 10 years, followed by 36.05% of patient who had the habit for 5-10 years, and this difference was found to be highly significant (p<0.01).

The study found that the most patients were diagnosed at stage III (69.96%), followed by stage II (21.03%). No cases of stage I were reported in the study population. Stage III and stage IV cancers were more common in males, accounting for 78.53% and 80.95%, respectively, while stage II was more prevalent in females (55.1%). Table No. 4 demonstrates that the association between gender and stage at diagnosis was highly significant (p<0.01). Furthermore, there was a strong association between the stage and age at diagnosis, with 69.96% of patients having stage III cancer, most of whom were in the 40-60 years age group, in contrast, stage IV cancer was predominantly found in individuals above 60 years (76.19%).

**Table No. 4:** Association of stage of oral cancer with gender and age at diagnosis:

| Gender | Stage at Diagnosis |  |  |  |  |  |
| --- | --- | --- | --- | --- | --- | --- |
|  | Stage I<sup>#</sup> | Stage II<sup>#</sup> | Stage III | Stage IV | Total | P value<0.01 |
| Male | 0 | 22(44.9%) | 128(78.53%) | 17(80.95%) | **167(71.67%)** |  |
| Female | 0 | 27(55.1%) | 35(21.47%) | 4(19.05%) | **66 (28.32%)** |  |
| **Total** | **0** | **49 (21.03%)** | **163 (69.96%)** | **21 (9.01%)** | **233 (100%)** |  |
| **Age at diagnosis (years)** |  |  |  |  |  |  |
| < 40 | **0** | 14(28.57%) | 4(2.45%) | 0 | **18(7.73%)** | **<0.01** |
| 40 – 60 | **0** | 32(65.30%) | 102(62.57%) | 5(23.80%) | **139(59.75%)** |  |
| > 60 | **0** | 3(6.12%) | 57(34.97%) | 16(76.19%) | **76(32.61%)** |  |
| **Total** | **0** | **49(21.03%)** | **163(69.96%)** | **21(9.01%)** | **233(100%)** |  |

## Discussion

In this study, out of 233 oral cancer patients, the highest proportion of cases were observed in individuals above 60 years of age, with the least affected group being those under 30 years. Most patients were male, resulting in a male-to-female ratio of 2.5:1. Among males, the most affected age group was above 60 years, while among females, it was 41-50 years, with no female patients below 40. These findings align with previous studies (5)(6)(12). However, Durgadevi Pancharethinam et al. reported a different pattern, with the most patients below 30 years of age and the minimum affected above 50 years (13). Regarding education, most oral cancer patients in our study were graduates, with 15.02% being illiterate, a finding consistent with Durgadevi Pancharethinam et al.’s study (13). Our results contrasted with another study where half of the patients had no schooling, and one-fourth had only primary education (5)(7). The distribution of oral cancer patients by occupation showed that the majority were clerical and skilled workers, with only 8.15% unemployed, in contrast to Ganesh R et al.’s findings, where a greater proportion of patients were unemployed and less skilled (5). As per Modified B.G. Prasad’s classification, revealed that most patients belonged to the lower-middle class, followed by the upper-lower class, with the least affected belonging to the upper class. These results were consistent with Akhilesh Krishna et al. and S P Khandekar et al.’s findings (12)(15), but Ganesh R et al. reported a higher proportion of upper-lower class patients (5).

The cheek was the most common site of oral cancer, followed by the tongue, while the lip was the least affected site. Both the floor of the mouth and other mouth parts were equally affected. These findings were consistent with previous studies (15)(16)(17)(18). Gender-wise distribution of cancer sites revealed that cheek and tongue were the most common sites in both males and females, with no significant association found between them, in agreement with a study conducted at a cancer hospital in Solapur (21). However, another study observed different results, with tongue and buccal mucosa being the most common sites in both genders (6). The distribution of patients by habit patterns showed that the tongue was the common site when a single habit, whether chewing, smoking, or alcohol consumption, was present. The cheek was the common site, in cases with a combination of more than one habit,. Patients with no habits were more likely to develop cancer in other mouth parts. These results aligned with Addala L et al. (6). However, Taban R J et al. reported that nontobacco users had more tumors of the buccal mucosa, tongue, and hard palate (23).

In this study, the majority of oral cancer patients had the habit of chewing tobacco alone, followed by tobacco chewing combined with smoking. Among males, multiple addictive habits were more common, while females predominantly had the habit of tobacco chewing alone. These findings were similar to other studies (7)(14)(19). The duration of habit revealed that most patients had the habit for more than 10 years, with a significant difference observed (p<0.01). The study also found that most patients were diagnosed at stage III, followed by stage II, and no cases of stage I were reported. This distribution differed by gender, with stage III and stage IV being more common in males and stage II more prevalent in females. These findings were highly significant (p<0.01). There was also a strong association between stage and age at diagnosis, with most stage III patients falling in the 40-60 years age group and stage IV patients primarily above 60 years.

## Conclusion

In this study, The male population, rural location, and additive habits mainly tobacco and alcohol are major risk factor for oral cancer. Males above 60 years and females in the 41-50 years age group were most affected by oral cancer. Cheek was the most common site for cancer, often associated with a single addictive habit, followed by the tongue, which was more commonly affected by a combination of two or more habits. Main focus on deaddiction and raising awareness for risk factors are essential steps to reduce the chances of developing oral cancer. Public health initiatives should emphasize education, prevention, and early treatment to combat addiction and oral cancer. There should be strict and legal provisions for selling tobacco products and alcohol.

## Conflict of Interest

None

## Funding

None

## Data Availability

All data produced in the present work are contained in the manuscript.

## Notes

### Competing Interest Statement

The authors have declared no competing interest.

### Funding Statement

This study did not receive any funding

### Author Declarations

S.S.Medical college, Rewa, Institutional Ethical Committee. No./IEC/MC/2020-470 Dated-08/01/2021 Approved

